# An analysis of COVID-19 article dissemination by Twitter compared to citation rates

**DOI:** 10.1101/2020.06.22.20137505

**Authors:** Nicholas Fabiano, Zachary Hallgrimson, Sakib Kazi, Jean-Paul Salameh, Stanley Wong, Abrar Kazi, Rudy R Unni, Ross Prager, Matthew DF McInnes

## Abstract

**Background:** The COVID-19 pandemic has resulted in over 1,000,000 cases across 181 countries worldwide. The global impact of COVID-19 has resulted in a surge of related research. Researchers have turned to social media platforms, namely Twitter, to disseminate their articles. The online database Altmetric is a tool which tracks the social media metrics of articles and is complementary to traditional, citation-based metrics. Citation-based metrics may fail to portray dissemination accurately, due to the lengthy publication process. Altmetrics are not subject to this time-lag, suggesting that they may be an effective marker of research dissemination during the COVID-19 pandemic.

**Objectives:** To assess the dissemination of COVID-19 articles as measured by Twitter dissemination, compared to traditional citation-based metrics, and determine article characteristics associated with tweet rates.

**Methods:** COVID-19 articles obtained from LitCovid published between January 1st to March 18th, 2020 were screened for inclusion. The following article characteristics were extracted independently, in single: Topic (General Info, Mechanism, Diagnosis, Transmission, Treatment, Prevention, Case Report, and Epidemic Forecasting), open access status (open access and subscription-based), continent of corresponding author (Asia, Australia, Africa, North America, South America, and Europe), tweets, and citations. A sign test was used to compare the tweet rate and citation rate per day. A negative binomial regression analysis was conducted to evaluate the association between tweet rate and article characteristics of interest.

**Results:** 1328 articles were included in the analysis. Tweet rates were found to be significantly higher than citation rates for COVID-19 articles, with a median tweet rate of 1.09 (IQR 6.83) tweets per day and median citation rate of 0.00 (IQR 0.00) citations per day, resulting in a median of differences of 1.09 (95% CI 0.86-1.33, *P* < .001). 2018 journal impact factors were positively correlated with tweet rate (*P* < .001). The topics Diagnosis (*P* = .01), Transmission (*P* < .001), Treatment (*P* = .01), and Epidemic Forecasting (*P* < .001) were positively correlated with tweet rate, relative to Case Report. The following continents of the corresponding author were negatively correlated with tweet rate, Africa (*P* < .001), Australia (*P* = .03), and South America (*P* < .001), relative to Asia. Open access journals were negatively correlated with tweet rate, relative to subscription-based journals (*P* < .001).

**Conclusions:** COVID-19 articles had significantly higher tweets rates compared to citation rates. This study further identified article characteristics that are correlated with the dissemination of articles on Twitter, such as 2018 journal impact factor, continent of the corresponding author, topic, and open access status. This highlights the importance of altmetrics in periods of rapidly expanding research, such as the COVID-19 pandemic to localize highly disseminated articles.

## Introduction

In December 2019, a collection of patients in Wuhan, China presented to hospital with respiratory symptoms and pneumonia. These patients were diagnosed with a novel coronavirus of the *Orthocoronavirinae* subfamily (SARS-CoV-2) [1,2]. SARS-CoV-2 is readily capable of transmission between human hosts, and is associated with a range of clinical severity from mild respiratory symptoms to severe hypoxemic respiratory failure and death [3,4]. Although originating in China, SARS-CoV-2 has resulted in over 1,000,000 cases of infection in 181 countries worldwide, and has claimed the lives of over 60,000 (as of April 5th) [5]. Whereas SARS-CoV-2 refers to the virus itself, its associated syndrome is referred to as coronavirus disease (COVID-19). COVID-19 has drastically impacted the lives of millions worldwide [5].

The global impact of COVID-19 has led to a surge of related research with over 1328 related articles indexed on PubMed as of March 20^th^, 2020 [6,7]. With the amount of research being performed, streamlined and timely methods for disseminating information to frontline clinicians has become central to providing evidence informed care. To facilitate this, clinicians have increasingly turned to social media platforms including Twitter, to share, discuss, and respond to peer reviewed studies, sometimes even before the formal peer review process [8].

The use of social media to disseminate knowledge among the medical community is not new, with Twitter serving as an important social media platform for scholarly communication [9,10]. Whereas traditional citation-based metrics can track the amount of times a publication is cited by other researchers, they do not consider the dissemination of the publication online across social media platforms. The online database Altmetric is a tool that tracks social media sharing, and then collects and displays metrics from sources including Twitter and Facebook [11,12]. The Altmetric database has previously been interrogated to explore how variables such as study discipline, number of authors, and country of origin influence the number of tweets pertaining to a given study [13,14] and as a tool to examine research dissemination. As there is a lack of correlation between altmetrics and citation rates, it has been suggested that twitter-based metrics reflect different, yet complementary information to traditionally used citation rates [15,16]. Therefore, altmetrics may serve as another tool to capture the wider impact of a given study [17].

Determining the impact and dissemination of research is especially important for the COVID-19 pandemic, as healthcare workers rely on current and impactful research to inform practice. Traditional citation metrics may fail to portray dissemination accurately, as there is a significant time-lag due to the publication process. Altmetrics do not have the same limitation, as they do not rely on the publication process, suggesting that they may provide an effective marker of research dissemination during the COVID-19 pandemic.

The objective of this study is to assess the impact of COVID-19 research articles as measured by Twitter dissemination, compared to a traditional citation-based metric. This study will also seek to determine which characteristics are associated with an article’s Twitter dissemination. This research serves as an important first step in understanding how best to rapidly disseminate new and important research during the COVID-19 pandemic.

## Methods

Research ethics board approval for this type of research is waived at our institution. The pre-specified study protocol is available on the Open Science Framework [18].

### Search strategy and selection

LitCovid [7], a curated literature hub that compiles research articles pertaining to COVID-19 on a daily basis, was searched on March 20th, 2020 for all publications included in the database to date. The search spanned from January 1^st^, 2020 to March 19^th^, 2020.

Two investigators (NF [second-year medical student] and ZH [third-year medical student]) independently screened each of the retrieved articles full-text for potential relevance. Articles were included if they discussed the novel SARS-CoV-2, or its associated syndrome COVID-19. Articles were excluded if they were retracted.

### Data collection

One investigator (ZH) extracted article title, authorship, publication date, continent of corresponding author, and topic from LitCovid. Two investigators (NF and SW [second year medical student]) extracted the 2018 journal impact factor and open access status from Clarivate Analytics Journal Citation Reports. Two investigators (SK [second year medical student], AK [fourth year mathematics student]) extracted the number of citations from the Web of Science (on March 23, 2020). One investigator (ZH) used Altmetric to derive the number of tweets made about an article (on March 23, 2020). Article DOIs were used to query the Altmetric Details Page API, version v1. If DOIs were not available, the article title was used to search in Altmetric Explorer. All extractions were done independently, in single.

Missing information was recorded as “not reported”. Default values were assigned in cases where the complete publication date was not available: in cases where the day of publication was unavailable, the first of the month was assigned as default. For journals not listed on Clarivate Analytics Journal Citation Reports or Web of Science, an impact factor of 0, and citation value of 0 were assigned, respectively. For articles with multiple topics or continents listed, the first listed topic or continent was taken.

### Data analysis

A sign test was used to compare the tweet rate and citation rate per day for each article. Significance was defined as p-value less than or equal to 0.05.

A negative binomial regression analysis was conducted to evaluate the association between tweet rate and article characteristics of interest: 2018 journal impact factors (median split), journal open access status (open access and subscription-based), topic (General Info, Mechanism, Diagnosis, Transmission, Treatment, Prevention, Epidemic Forecasting, and Case Report), and continent of corresponding author (Africa, Australia, Europe, North America, South America, and Asia), starting from the date of publication to March 23rd, 2020. Positive and negative regression coefficients were calculated to evaluate the associations between a specific article characteristic of interest and tweet rate. Predictive Mean Matching imputation was performed for 3 variables (open access status, continent of corresponding author, and topic of article) with a total missingness proportion of 20% out of the 1328 included articles. Imputation was performed using the MICE package in R [19–22].

Using R software for analysis (version 3.6.3, R Foundation for Statistical Computing), the p-values for the association of the variables of interest with tweet rate were obtained by means of likelihood ratio testing with a chi-squared mode, where a value of 0.05 was used as the threshold for significance.

## Results

### Article selection

The search of the LitCovid database yielded 1328 articles. All articles fulfilled the inclusion and exclusion criteria.

### Article characteristics

The included article characteristics are presented in Table 1. Publication dates ranged from January 1^st^, 2020 to March 19^th^, 2020. Exact publication dates were available for 1314 articles (98.94%) and default days were assigned to 14 articles (1.05%). On the LitCovid database, a single topic was listed for 1180 articles (88.86%) and multiple topics were listed for 148 articles (11.14%). A single continent of the corresponding author was listed for 1291 articles (97.21%) and multiple were listed for 37 articles (2.79%). The included articles were published in 326 journals, with a median impact factor of 3.73.

**Table 1:**
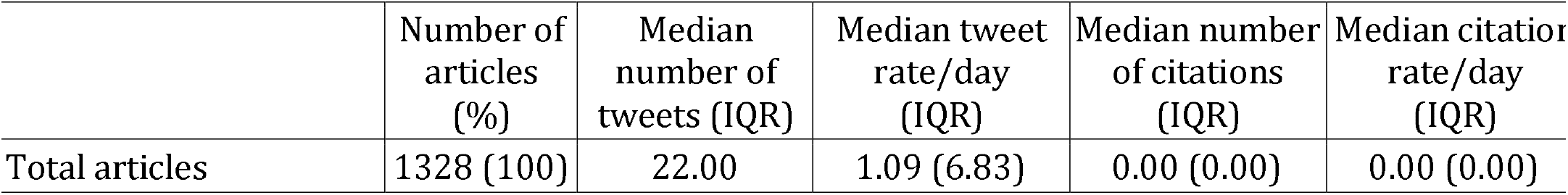
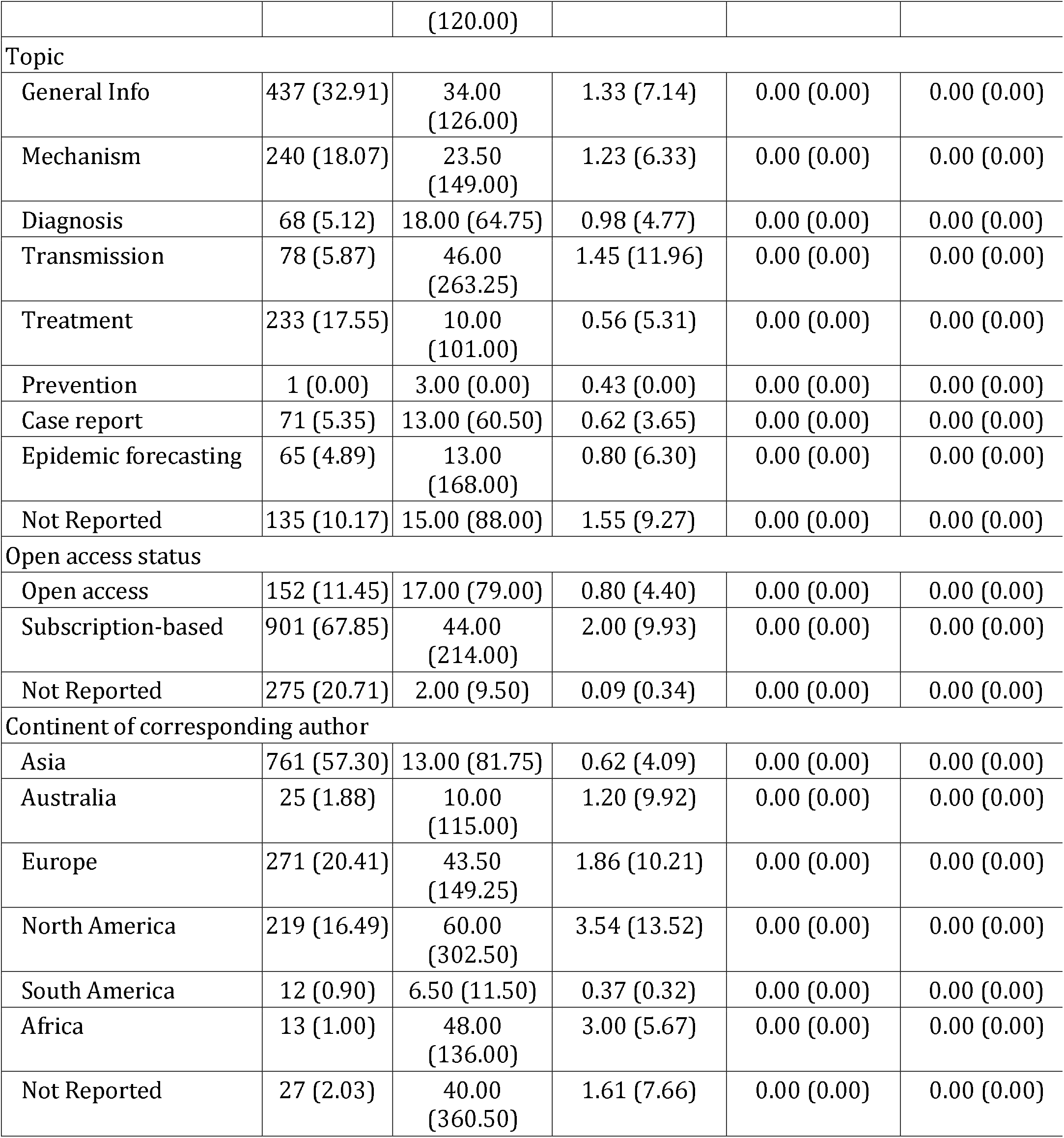
Summary characteristics of included articles.

### Comparison of dissemination: tweet rates versus citation rates

Tweet rates were found to be significantly higher than citation rates for COVID-19 articles, with a median tweet rate of 1.09 (IQR 6.83) tweets per day and median citation rate of 0.00 (IQR 0.00) citations per day. The tweet rate ranged from 0 to 2652.67 tweets per day and the citation rate ranged from 0 to 1.78 citations per day. The resulting median of differences was 1.09 (95% CI 0.86-1.33, *P* < .001).

### Association of Twitter dissemination with article characteristics

A positive association between 2018 impact factor and tweet rate was present. Assessment of the association between topic and tweet rate yielded a positive association present for the topics Diagnosis, Epidemic Forecasting, Transmission, and Treatment, relative to Case Reports, with the largest association observed in Epidemic Forecasting (regression coefficient: 1.95, *P* < .001). A negative association was found between open access journals and tweet rate, relative to subscription-based journals. Further, analysis of the association between continent of corresponding author and tweet rate found a negative association for articles published in Australia, Africa, and South America, relative to those published in Asia. A significant negative association was found between 2018 journal impact factor and open access status. Regression coefficients for the article characteristics of interest are presented in Table 2.

**Table 2:** Negative binomial regression model evaluating the association between tweet rate and 2018 journal impact factor, open access status, topic, and continent of corresponding. Variable categories with coefficients indicated as “REF” were used as the reference category for their respective categorical variables. Significance defined as *P* < .05

| Variable | Regression coefficient | Standard error | p-value |
| --- | --- | --- | --- |
| Intercept | 1.42 | 0.24 | <.001 |
| 2018 impact factor | 2.33 | 0.13 | <.001 |
| Open access status |  |  |  |
| Open access | -1.19 | 0.19 | <.001 |
| Subscription-based | REF | N/A | N/A |
| 2018 impact factor X<br>Open access | -0.44 | 0.33 | .03 |
| Topic |  |  |  |
| General Info | 0.32 | 0.24 | .19 |
| Mechanism | 0.41 | 0.25 | .11 |
| Diagnosis | 0.85 | 0.30 | .01 |
| Transmission | 1.08 | 0.30 | <.001 |
| Treatment | 0.71 | 0.25 | .01 |
| Prevention | -2.17 | 2.05 | .29 |
| Epidemic Forecasting | 1.95 | 0.39 | <.001 |
| Case Report | REF | N/A | N/A |
| Continent of corresponding author |  |  |  |
| Africa | -2.18 | 0.53 | <.001 |
| Australia | -0.87 | 0.39 | .03 |
| Europe | -0.11 | 0.14 | .46 |
| North America | -0.15 | 0.15 | .34 |
| South America | -2.85 | 0.58 | <.001 |
| Asia | REF | N/A | N/A |

## Discussion

The objective of this study was to compare tweet rates to citation rates for articles about COVID-19 that were extracted from the LitCovid database, and to identify article characteristics that correlate with the number of tweets an article receives. The tweet rates were found to be significantly higher that the citations rates. The number of tweets an article received was positively correlated with 2018 journal impact factor and topic (Diagnosis, Transmission, Treatment, and Epidemic Forecasting relative to Case Report) and was negatively correlated with open access journals (relative to subscription-based journals) and the continent of the corresponding author (Africa, Australia, and South America, relative to Asia).

A significant difference was found between the tweet and citation rate. This result is in accordance with previously described literature from Haustein et. al. which found a lack of association between tweets [15] and citation rates for journal articles in the biomedical sciences and from Costas et. al. showing a weak correlation between tweets and citation rates from different disciplines indexed from Web of Science [16]. Thus, altmetrics provide different and supplementary information to previously used citation based metrics for article dissemination analysis. Further, the lack of association demonstrates that during the COVID-19 pandemic, citation rates are not optimal to quantify article dissemination due to their associated time-lag. As altmetrics do not suffer from this limitation, they are better equipped to quantify dissemination of articles and provide an objective metric for healthcare workers or researchers to find impactful research, as evidenced by our finding that tweet rates were significantly higher than citation.

A journal’s 2018 impact factor was found to be positively associated with tweet rate. This is fitting as journals with higher impact factors are often exposed to larger audiences, increasing the likelihood that articles published in these journals would be tweeted about.

The continent of the corresponding author was found to impact the number of tweets with South America having the largest negative association, followed by Africa, then Australia, relative to articles published from Asia. An analysis of worldwide Twitter usage using tweet geolocation, found that Twitter has a strong prevalence in North America and Europe compared to other continents with African and South American countries having the lowest involvement [23]. This could explain the lower number of tweets associated with Africa and South America. There was also a lower research output, as reflected by the relatively low number of articles from Africa, South America and Australia which could account for the lower number of tweets received. Similarly, Asia had the highest number of articles, followed by Europe and North America which could account for their higher number of tweets. The number of tweets also followed the progression of COVID-19 across continents, with early cases surging in Asia, followed by Europe, then North America. Therefore, when looking at articles on social media platforms it is important to acknowledge that the articles presented may be biased towards countries with higher social media use or in general more research on the topic.

Epidemic forecasting, Transmission, and Diagnosis had significantly higher rates of tweets (compared to General Info, Mechanism, and Prevention), relative to Case Reports. This result could convey an increased interest in these topics for the COVID-19 pandemic, resulting in increased tweet rates. Interestingly, these topics had a relatively low amount of articles, relative to the other topics, suggesting that general perceived interest in a topic and not absolute number of publications could influence online dissemination.

Surprisingly, journals that were open access had a negative association of tweets compared to subscription-based journals. This finding was supported by the statistically significant interaction between 2018 impact factor and open access status in the regression model. Since journals with higher impact factor were more likely to be subscription-based, articles published in subscription-based journals had an increased likelihood of Twitter dissemination. It is important to note that many prominent subscription-based journals have made COVID-19 related research open access in an effort to improve dissemination to readers. This could have influenced the number of tweets made about articles from subscription-based journals.

There are a few limitations to this study. There may be residual confounding variables that were not measured in this analysis. The authors acknowledge that self-promotion (eg: authors tweeting out their own article) may influence the number of tweets an article receives; however, the extent is difficult to quantify. Further, this study did not quantify the sentiment of the tweets and citations, which could provide useful information. However, it has been found that 94.8% of tweets linking to scientific papers and 75.85% of citations in clinical trial papers were neutral [24,25]. Thus, this suggests that the majority of citations and tweets within this study were neutral in sentiment. Further research should be conducted in this area to clarify this. Also, there is a loss in precision due to the use of categorical variables such as the use of continents instead of countries and use of impact factor median split instead of impact factor. This was used in order to get an appropriate fit of the model, but loss of precision is a consequence. Similarly, the relatively high proportion of missing values (20%) could have impacted the robustness of the model fit to the data, despite the imputation performed to address this concern.

Research surrounding the COVID-19 pandemic is being performed at an ever increasing rate. This study represents a snapshot of the early research into the pandemic, repeating this analysis as more research becomes available will be important to confirm findings. In addition, sentiment analysis (categorizing the content of the tweets as positive, negative, or neutral) may be valuable to gain further insight into the content of tweets shared online for a given article. A tool known as SentiStrength has shown potential in this area by categorizing sentiment strengths of social media text, and may prove useful in this type of analysis [26].

LitCovid provides a centralized curated hub for readers to find research related to COVID-19. However, not all periods of rapidly expanding research has this same easily, accessible resource. Thus, the results of this study suggest that the amount of tweets, or online dissemination in general by means of altmetrics, can be used to locate highly disseminated articles in other areas of rapidly evolving research, such as the vaping-associated lung disease which sporadically appeared in the summer of 2019 [27], where citation based metrics may fail to capture article dissemination.

## Conclusion

This study collected data from LitCovid on articles related to COVID-19 and showed that these articles had significantly higher tweets rates compared to citation rates. This study also identified that the number of tweets an article received was positively correlated with 2018 journal impact factor and topic (Diagnosis, Transmission, Treatment, and Epidemic Forecasting relative to Case Report) and was negatively correlated with open access journals (relative to subscription-based journals) and the continent of the corresponding author (Africa, Australia, and South America, relative to Asia). This analysis expands the current literature on the use of altmetrics in categorizing research dissemination and its practical use in periods of rapidly expanding research, such as the COVID-19 pandemic, to localize highly disseminated articles.

## Data Availability

All articles were extracted from the LitCovid database. Data is available upon request.

https://www.ncbi.nlm.nih.gov/research/coronavirus/

## References

1. Zhu N, Zhang D, Wang W, Li X, Yang B, Song J, Zhao X, Huang B, Shi W, Lu R, Niu P, Zhan F, Ma X, Wang D, Xu W, Wu G, Gao GF, Tan W. A Novel Coronavirus from Patients with Pneumonia in China, 2019. N Engl J Med; 2020 Feb 20;382(8):727–733. PMID:31978945

2. Gorbalenya AE, Baker SC, Baric RS, de Groot RJ, Drosten C, Gulyaeva AA, Haagmans BL, Lauber C, Leontovich AM, Neuman BW, Penzar D, Perlman S, Poon LLM, Samborskiy DV, Sidorov IA, Sola I, Ziebuhr J, Coronaviridae Study Group of the International Committee on Taxonomy of Viruses. The species Severe acute respiratory syndrome-related coronavirus11: classifying 2019-nCoV and naming it SARS-CoV-2. Nat Microbiol Nature Publishing Group; 2020 Apr;5(4):536–544. [doi: 10.1038/s41564-020-0695-z]

3. Guan W, Ni Z, Hu Y, Liang W, Ou C, He J, Liu L, Shan H, Lei C, Hui DSC, D. B, Li L, Zeng G, Yuen K-Y, Chen R, Tang C, Wang T, Chen P, Xiang J, Li S, Wang J, Liang Z, Peng Y, Wei L, Liu Y, Hu Y, Peng P, Wang J, Liu J, Chen Z, Li G, Zheng Z, Qiu S, Luo J, Ye C, Zhu S, Zhong N. Clinical Characteristics of Coronavirus Disease 2019 in China. N Engl J Med; 2020 Feb 28;0(0):null. PMID:32109013

4. Arentz M, Yim E, Klaff L, Lokhandwala S, Riedo FX, Chong M, Lee M. Characteristics and Outcomes of 21 Critically Ill Patients With COVID-19 in Washington State. JAMA [Internet] 2020 Mar 19 [cited 2020 Mar 27]; PMID:32191259

5. Coronavirus latest: confirmed cases cross the one-million mark. Nature [Internet] Nature Publishing Group; 2020 Apr 1 [cited 2020 Apr 4]; PMID:32152592

6. Callaway E, Cyranoski D, Mallapaty S, Stoye E, Tollefson J. The coronavirus pandemic in five powerful charts. Nature [Internet] Nature Publishing Group; 2020 Mar 18 [cited 2020 Mar 21]; PMID:32203366

7. Chen Q, Allot A, Lu Z. Keep up with the latest coronavirus research. Nature Nature Publishing Group; 2020 Mar 10;579(7798):193–193. PMID:32157233

8. KupferschmidtFeb. 26 K, 2020, Pm 2:05. ‘A completely new culture of doing research.’ Coronavirus outbreak changes how scientists communicate [Internet]. Sci AAAS. 2020 [cited 2020 Mar 21]. Available from: https://www.sciencemag.org/news/2020/02/completely-new-culture-doing-research-coronavirus-outbreak-changes-how-scientists

9. Sugimoto CR, Work S, Larivière V, Haustein S. Scholarly use of social media and altmetrics: A review of the literature. J Assoc Inf Sci Technol 2017;68(9):2037–2062. [doi: 10.1002/asi.23833]

10. Rowlands I, Nicholas D, Russell B, Canty N, Watkinson A. Social media use in the research workflow. Learn Publ 2011;24(3):183–195. [doi: 10.1087/20110306]

11. Altmetric.com. “Knowledge Base”. http://support.altmetric.com/knowledgebase.

12. Adie E, Roe W. Altmetric: enriching scholarly content with article-level discussion and metrics. Learn Publ 2013;26(1):11–17. [doi: 10.1087/20130103]

13. Andersen JP, Haustein S. Influence of Study Type on Twitter Activity for Medical Research Papers. In: Salah AA, Tonta Y, Salah A a. A, Sugimoto C, Al U, editors. Proc Issi 2015 Istanb 15th Int Soc Scientometr Informetr Conf Leuven: Int Soc Scientometrics & Informetrics-Issi; 2015. p. 26–36.

14. Haustein S. Scholarly Twitter Metrics. In: Glänzel W, Moed HF, Schmoch U, Thelwall M, editors. Springer Handb Sci Technol Indic [Internet] Cham: Springer International Publishing; 2019 [cited 2020 Mar 21]. p. 729–760. [doi: 10.1007/978-3-030-02511-3_28]

15. Haustein S, Peters I, Sugimoto CR, Thelwall M, Larivière V. Tweeting biomedicine: An analysis of tweets and citations in the biomedical literature. J Assoc Inf Sci Technol 2014;65(4):656–669. [doi: 10.1002/asi.23101]

16. Costas R, Zahedi Z, Wouters P. Do “altmetrics” correlate with citations? Extensive comparison of altmetric indicators with citations from a multidisciplinary perspective. J Assoc Inf Sci Technol 2015;66(10):2003–2019. [doi: 10.1002/asi.23309]

17. Hall N. The Kardashian index: a measure of discrepant social media profile for scientists. Genome Biol 2014 Jul 30;15(7):424. PMID:25315513

18. Hallgrimson Z, Fabiano N, Kazi S, Wong S, Kazi A, Unni R, Prager R, McInnes M. Altmetrics of COVID-19 research: An analysis of study dissemination by Twitter compared to citation rates. OSF; 2020 Mar 27 [cited 2020 Apr 4]; [doi: None]

19. Missing-Data Adjustments in Large Surveys: Journal of Business & Economic Statistics: Vol 6, No 3 [Internet]. [cited 2020 Apr 4]. Available from: https://amstat.tandfonline.com/doi/abs/10.1080/07350015.1988.10509663

20. Morris TP, White IR, Royston P. Tuning multiple imputation by predictive mean matching and local residual draws. BMC Med Res Methodol 2014 Jun 5;14(1):75. PMID:24903709

21. Buuren S van. Flexible Imputation of Missing Data, Second Edition. CRC Press; 2018. ISBN:978-0-429-96034-5

22. Buuren S van, Groothuis-Oudshoorn K. mice: Multivariate Imputation by Chained Equations in R. J Stat Softw 2011 Dec 12;45(1):1–67. [doi: 10.18637/jss.v045.i03]

23. Leetaru K, Wang S, Padmanabhan A, Shook E. Mapping the global Twitter heartbeat: The geography of Twitter. First Monday [Internet] 2013 May 6 [cited 2020 Mar 31];18(5). [doi: 10.5210/fm.v18i5.4366]

24. Friedrich N, Bowman TD, Stock WG, Haustein S. Adapting sentiment analysis for tweets linking to scientific papers. In: Salah AA, Tonta Y, Salah A a. A, Sugimoto C, Al U, editors. Proc Issi 2015 Istanb 15th Int Soc Scientometr Informetr Conf Leuven: Int Soc Scientometrics & Informetrics-Issi; 2015. p. 107–108.

25. Xu J, Zhang Y, Wu Y, Wang J, Dong X, Xu H. Citation Sentiment Analysis in Clinical Trial Papers. AMIA Annu Symp Proc AMIA Symp 2015;2015:1334–1341. PMID:26958274

26. Thelwall M, Buckley K, Paltoglou G, Cai D, Kappas A. Sentiment strength detection in short informal text. J Am Soc Inf Sci Technol 2010;61(12):2544–2558. [doi: 10.1002/asi.21416]

27. Layden JE, Ghinai I, Pray I, Kimball A, Layer M, Tenforde MW, Navon L, Hoots B, Salvatore PP, Elderbrook M, Haupt T, Kanne J, Patel MT, Saathoff-Huber L, King BA, Schier JG, Mikosz CA, Meiman J. Pulmonary Illness Related to E-Cigarette Use in Illinois and Wisconsin — Final Report. N Engl J Med; 2020 Mar 5;382(10):903–916. [doi: 10.1056/NEJMoa1911614]

